# Phased implementation of COVID-19 vaccination: rapid assessment of policy adoption, reach and effectiveness to protect the most vulnerable in the US

**DOI:** 10.1101/2021.02.19.21252118

**Authors:** Yun Li, Moming Li, Megan Rice, Yanfang Su, Chaowei Yang

## Abstract

**Objectives:** The US and rest of the world have suffered from the COVID-19 pandemic for over a year. The high transmissibility and severity of this virus have provoked governments to adopt a variety of mitigation strategies. Some of these previous measures, such as social distancing and mask mandates, were effective in reducing the case growth rate yet became economically and administratively difficult to enforce as the pandemic continued. In late December 2020, COVID-19 vaccines were first approved in the US and states began a phased implementation of COVID-19 vaccination. However, there is limited quantitative evidence regarding the effectiveness of the phased COVID-19 vaccination. This study aims to provide a rapid assessment of the adoption, reach, and effectiveness of the phased implementation of COVID-19 vaccination.

**Methods:** We utilize an event-study analysis to evaluate the effect of vaccination on the state-level, daily COVID-19 case growth rate.

**Results:** Through this analysis, we assert that vaccination is effective in reducing the spread of COVID-19 shortly after the first shots were given. Specifically, the case growth rate declined by 0.124, 0.347, 0.345, 0.464, 0.490, and 0.756 percentage points corresponding to the 1-5, 6-10, 11-15, 16-20, 21-25, and 26 or more day periods after the initial shots.

**Conclusions:** The findings could be insightful for policymakers as they work to optimize vaccine distribution in later phases, and also for the public as the COVID-19 related health risk is a contentious issue.

## Introduction

In late December 2019, a pneumonia-like illness was reported in Wuhan, China, which came to be known as COVID-19. In the US, the first case was documented in late January in Washington state. By mid-March, all 50 states had reported cases and by mid-April the US had the most COVID-19 deaths in the world (Kantis et al., 2020). To combat the rapidly growing pandemic, many states implemented lockdown measures which coincided with an economic downturn (IMF, 2020). Therefore, despite growing cases, lockdowns were eased in many states by April (Washington Post Staff, 2020). Additionally, there has been a decline in compliance towards social distancing measures which are often voluntary and hard to enforce (Boseley, 2020). As lockdowns are generally not economically sustainable, alternative methods to control the pandemic have received growing attention. Accordingly, many research groups have worked on developing a COVID-19 vaccine. The Centers for Disease Control and Prevention (CDC) has stated that vaccines are vital in tandem with other social distancing measures. In the US, many companies started developing COVID-19 vaccines in March (HHS, 2020). In December, the Food and Drug Administration (FDA) authorized the emergency use of both the Pfizer-BioNTech and Moderna vaccines. Despite their notoriously rapid development, clinical trials have found both vaccines to be more than 90% effective with only a few, minor side effects (Luthi, 2020).

Once the vaccine efficacy was thoroughly tested, the government started planning distribution efforts through a program known as Operation Warp Speed (HHS, 2020). The first round of vaccines was delivered to states, territories, and some federal agencies in late December (Silberner, 2020; Ivory et al., 2021). Ultimately, the states are in charge of the local administration of vaccines. However, as the supply is currently limited the Advisory Committee on Immunization Practices (ACIP) has suggested taking a phased approach to vaccination (CDC, 2020). This approach was made in an attempt to decrease deaths and serious illnesses while preserving a functioning society and reducing the burden on those who have been disproportionately harmed by the virus (CDC, 2021). The federal government anticipates vaccinating 100 million people in the US by the end of February 2021. However, the vaccine rollout has been slow (Silberner, 2020) likely due to the limited supply and inefficient distribution (CDC, 2020; Robbins et al., 2020; Romero et al., 2021). It is important to rapidly assess the phased vaccination approach in terms of adoption, reach, and effectiveness, despite this low coverage, in order to better plan for vaccination later on. Indeed, an evaluation of the pandemic in California near the end of January 2021 noted that cases (The COVID Tracking Project, 2021) and hospitalizations were starting to decline at the same time vaccination coverage was increasing (Los Angeles Times Staff, 2021).

Similar to social distancing policies, the COVID-19 vaccination has become a contentious issue especially as there is limited direct evidence regarding the effectiveness of vaccination. Many people who protested early pandemic measures, such as stay at home orders and mask mandates, are likewise protesting mandatory vaccines (Luthi, 2020). This opposition may be due to firm beliefs in medical autonomy, vaccine conspiracy theories (Ball, 2020), concerns about vaccine side effects, mistrust of the government (Dwyer, 2020), and mistrust of healthcare (Coustasse et al., 2020). However, COVID-19 vaccine opinions and knowledge have varied overtime. There has been a noticeable increase in the public’s willingness to take the vaccine as it has moved into later stages of development (Luthi, 2020; Dwyer, 2020). Increasing the public trust in COVID-19 vaccines is critical as many experts predict that a vaccination rate between 70% and 80% is needed for herd immunity (Maragakis, 2021). As manufacturing and distribution become more systematic, it will become more important to eliminate other barriers to widespread vaccination efforts such as public opposition.

In order to increase the public’s willingness to take the vaccine, some studies have recommended utilizing public information campaigns, financial investments, and other measures to highlight the effectiveness, benefits, and safety of the vaccine (Paltiel et al., 2020). Furthermore, other studies have emphasized the importance of making sure these campaigns receive high coverage and are spread to the public quickly, as the pandemic is an urgent issue (Buonomo et al., 2020). Without sufficient knowledge, it is often hard to persuade people to accept recommendations (Wolfson, 2020). For example, mask mandates ultimately gained more widespread acceptance once thorough research highlighted their success (Lyu et al., 2020). Indeed, as the vaccine supply increases, distribution will move towards phases that include a broader range of people who are eligible for vaccination (CDC, 2020). Therefore, evaluating the effectiveness of vaccination in controlling the spread of COVID-19 is important to ensure a smooth transition between phases by increasing vaccination acceptance and coverage.

Currently, there is limited research done to quantitatively evaluate the effectiveness of the phased COVID-19 vaccination approach. Through a natural experiment, we investigate how statewide vaccination efforts affect the spread of COVID-19 in the US during the early stages of vaccination. Specifically, we use an event-study analysis to estimate the effects of state vaccinations on the state-level confirmed case growth rate over different time periods. Vaccination may play a vital role as states are reopening and social distancing policies are becoming less stringent (Boseley, 2020; Hills, 2021). It is necessary to do a rapid assessment of vaccination effectiveness during the initial stages of implementation as concrete evidence of success and safety is important to inform policymakers who direct administration efforts, and the public in order to achieve widespread vaccination.

## Study area and data

### Vaccination Data

Records detailing when the COVID-19 vaccine was first administered in every state were gathered in this study by searching and reviewing the public news and governmental pages. Our study focuses on state-level COVID-19 vaccination beginning with the first shot which was given in New York on December 14, 2020. Following the CDC guidelines, frontline healthcare workers and long-term care facility staff and residents were given the highest priority in receiving the vaccine in phase one (CDC, 2020). However, the vaccination process, including the definition of groups given priority, have varied among the states. Detailed information on the adoption of vaccination in different US states can be found in the result section. In addition, we collected vaccination data from October 1, 2020 to January 26, 2021. A repository shared by the Johns Hopkins University Centers for Civic Impact COVID-19 data analysis and visualizations (https://github.com/govex/COVID-19) contains state-level reports which record the daily cumulative number of doses administered to the public. Specifically, we divided the cumulative number of doses administered by a state’s population size to find the daily ratio of doses administered.

### State-Level Policy Data

Statewide policy data was produced by the NSF spatiotemporal innovation center based on the Oxford COVID-19 Government Response Tracker project schema (Hale et al., 2020). This dataset describes common measures governments have taken in the fight against COVID-19 and their implementation overtime. These policies include school closures, workplace closures, the cancellation of public events, restrictions on gatherings, public transport closures, stay at home orders, internal movement restrictions, and international travel controls. The extent of government action is reflected using a stringency index ranging from 0 to 100; the larger the value, the more stringent a certain policy measure is. The entire dataset can be found on Github (https://github.com/OxCGRT/covid-policy-tracker). A lot of research has found that social distancing policies and mask mandates affect the growth rate of COVID-19 cases (Courtemanche et al., 2020; Li et al., 2020; Lyu et al., 2020), thus, the stringency index was included in this study to control for the effects of policy stringency during the study period.

### State-level confirmed case data

The NSF spatiotemporal center provided a data repository detailing daily COVID-19 data (Sha et al., 2020; Yang et al., 2020). Specifically, our study used the US state-level confirmed case data. This level of data was used as most vaccination plans were designed and implemented by individual states (Silberner, 2020; Ivory et al., 2021). Only confirmed case data was used in this research since we focused on the impact of vaccines on the spread of COVID-19. The daily confirmed case data was gathered from this Github repository, (https://github.com/stccenter/COVID-19-Data/tree/master/US), and daily growth rates of confirmed cases were derived from the original dataset as introduced in the methodology section.

## Methodology

### Data processing

#### (1) Vaccination

State-level vaccination data was encoded as binary data, with 0 indicating that the COVID-19 vaccine had not been administered in a certain state and 1 indicating that people had started to take the vaccine in that state. To assess the vaccination effects, a reference group representing the 1-5 days before the start of vaccination was selected. Additionally, we analyzed the effects of the vaccination over six post-periods (1-5 days, 6-10 days, 11-15 days, 16-20 days, 21-25 days, and 26+ days)—which represented days after the first shot of the COVID-19 vaccine was administered, and four pre-event trends (6-10 days, 11-15 days, 16-20 days, and 20+ days)—which represented days before the vaccination started. Multiple 0-1 indicators were generated to indicate these pre- and post-periods. Each day in the analytical period was assigned with one indicator by comparing t to the time range each indicator represented. Here, t is the cumulative number of days since the first shot of the vaccine was administered in a certain state.

#### (2) Confirmed case growth rate

To estimate the effect of vaccination, we calculated the daily, state-level confirmed case growth rate, which is the difference between the natural logarithms of the cumulative counts of confirmed cases on a given day and that of the prior day. This difference was then multiplied by 100 to enable the daily growth rate to be given in percentage points and so that the estimated regression coefficients could be interpreted as percentage point changes in the growth rate.

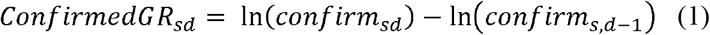

### Statistical analysis

In this study, we employed a fixed effects panel regression model incorporating the event-study design to examine the effects of statewide phased COVID-19 vaccination on the spread of COVID-19 in the US. The differences between the availability of vaccines before and after the initial vaccination in the states enabled estimation in the context of a natural experiment. We examined if the trend in the COVID-19 confirmed case growth rate was differentiated before and after the start of vaccination. The response variable in the model was the confirmed case growth rate as defined in Eq (1). The effect of periods of interest was defined in the Vaccination section (the six post periods after the first shot of the COVID-19 vaccine were coded *i* = 1,…, 6, the four pre-event periods were coded *i* = −4,…, −1, and the reference period was *i* = 0). In addition to the differences in vaccination efforts, we also included the test growth rate, the policy stringency index (Hale et al., 2020), and the dose administrative ratio as control variables.

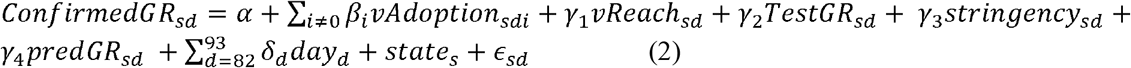

In the above regression specification, *νAdoption*_*sdi*_ denotes the indicator variable for the policy of vaccination belonging to the period *i* for state s and day d (normalized to 0), *νReach*_*sd*_ denotes the dose administrative ratio for state *s* and day *d*. Additionally, state*s* denotes the state fixed effects; *confrimedGR*_*sd*_, *TestGR*_*sd*_, and *stringency*_*sd*_ denote, respectively, the confirmed case growth rate, the test case growth rate, and the stringency index, for state *s* and day *d*.

Meanwhile, since the first shot of the vaccine was administered on December 14 and most states started vaccination in the following few days, the effects of vaccination may be affected by the coinciding holiday season. In the regression, we added holiday fixed effects to control for possible confounding brought by the Christmas holiday. We denote the holiday season indicator by *day*_*d*_, where *d* =82 and *d* = 93 corresponds to 7 days before and after Christmas. To capture the pre-vaccination trends, a linear calendar day trend from October 31 to December 13 was estimated using a fixed effects panel regression model for the confirmed case growth rate. Then, daily confirmed case growth rates were predicted for each state and for the whole time period based on the estimated pre-trend. The predicted case growth rates for state s and day *d*, denoted by *predGR*_*sd*_, were included as a covariate in the regression model to control for the effects of the pre-vaccination period. Respecting the clustered nature of the repeated measurements in the panel dataset, we calculated robust standard errors clustered by states.

## Result

### Vaccination policy adoption

Vaccination efforts were guided at the state level. All states started the vaccination of priority groups by December 16; however, some states initiated their vaccine administration quicker than others. The collected information shows that about half of all states administered their first shot of the COVID-19 vaccine on December 14, 2020, with the overall first shot in the US being given in New York on the same day. Most other states started to vaccinate priority groups in the following days. Detailed information of the dates and news sources can be found in the appendix.

The delivery of vaccinations to the public was implemented in settings such as healthcare facilities, stadiums, and pharmacies, which received doses from the state (Gold, 2021). Since the supply of vaccines is limited, the ACIP suggests offering vaccination in a phased approach (CDC, 2020). This approach gives vulnerable populations high priority. According to the recommendation, Phase 1a includes healthcare personnel and long-term care facility residents.

Phase 1b includes persons ≥75 years of age and frontline essential workers. Phase 1c includes persons 65-74 years of age, persons 16-64 years of age with high-risk medical conditions, and other essential workers. However, as distribution was left up to individual states, many phases were defined slightly differently. The CDC phase definition that most closely aligned to the state’s most recently vaccinated populations (as of January 26, 2021) was used to categorize the state in Table 2, which represents the state-level implementation of vaccination. Information regarding each state’s specific vaccination phases were gathered from USAFacts (https://usafacts.org/visualizations/covid-vaccine-tracker-states/#). The states also show variation in their vaccination progress. Although all states are still in phase 1 as of January 26, some states have already entered phase 1c while others are still in phase 1a.

**Table 2:** The implementation status of COVID-19 vaccination in every state (as of January 26, 2021)

| Phase | States in a Certain Phase |
| --- | --- |
| Phase 1a | Arizona, Florida, Georgia, Hawaii, Kentucky, Maine, Oregon, Utah |
| Phase 1b | Arkansas, California, Colorado, Delaware, Idaho, Illinois, Indiana, Iowa, Kansas, Louisiana, Massachusetts, Michigan, Montana, New Mexico, North Carolina, North Dakota, Ohio, Oklahoma, Rhode Island, South Carolina, Tennessee, Vermont, Washington, Wyoming |
| Phase 1c | Alabama, Alaska, Connecticut, Maryland, Minnesota, Mississippi, Missouri, Nebraska, Nevada, New Hampshire, New Jersey, New York, Pennsylvania, South Dakota, Texas, Virginia, West Virginia, Wisconsin |

### Vaccination reach

Figure 1 presents the cumulative number of administered doses in each state by certain dates during the analysis period. Throughout this time, some states saw rapid growth in the number of doses they administered. For example, Texas and Florida slowly started vaccination on 2020-12-14, as was the case with most other states during the start of vaccination efforts. However, by 2021-01-26, Florida and Texas had some of the highest totals of administered doses. Many states did not see such a significant change throughout the analysis period. Arkansas, for example, had low numbers of administered doses during the whole period. Overtime, Texas and Florida likely became more efficient in their vaccination efforts.

**Figure 1:**
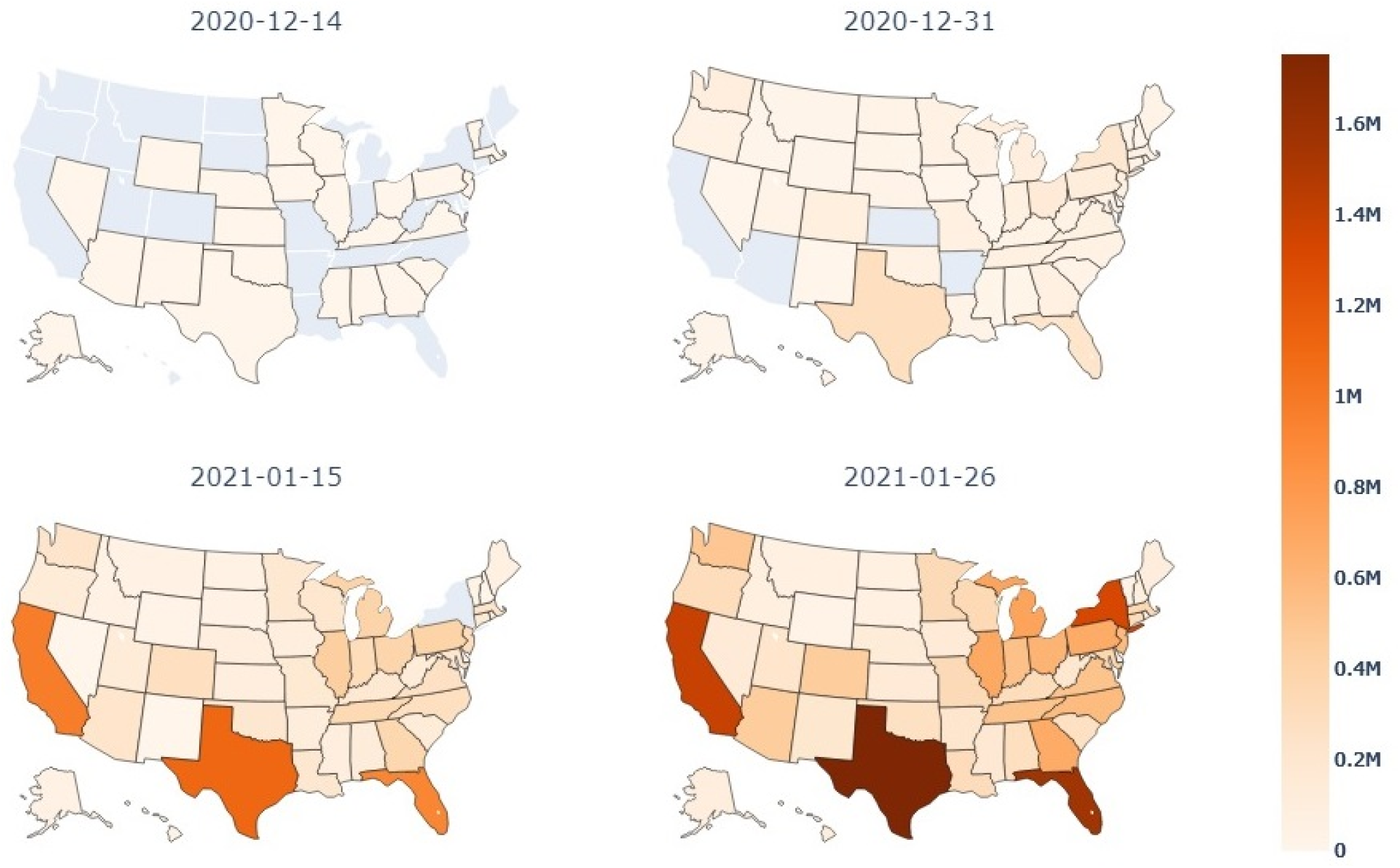
Vaccination doses administered in the study period.

The attempts of each state to reach the public can also be seen in Figure 2, which shows the percentage of the population, in each US state who received at least 1 vaccine dose by the same four dates described above. Throughout the analysis period, Texas and Florida were able to notably increase the percentage of their populations who received at least 1 dose of the vaccine. West Virginia also saw large increases in this population percentage and has been commended for its vaccination efforts (Anderson et al., 2021). In terms of vaccinations, some states have been more effective in reaching the public as seen by changes in the number of doses administered and the percent of the population with at least 1 vaccine dose overtime.

**Figure 2:**
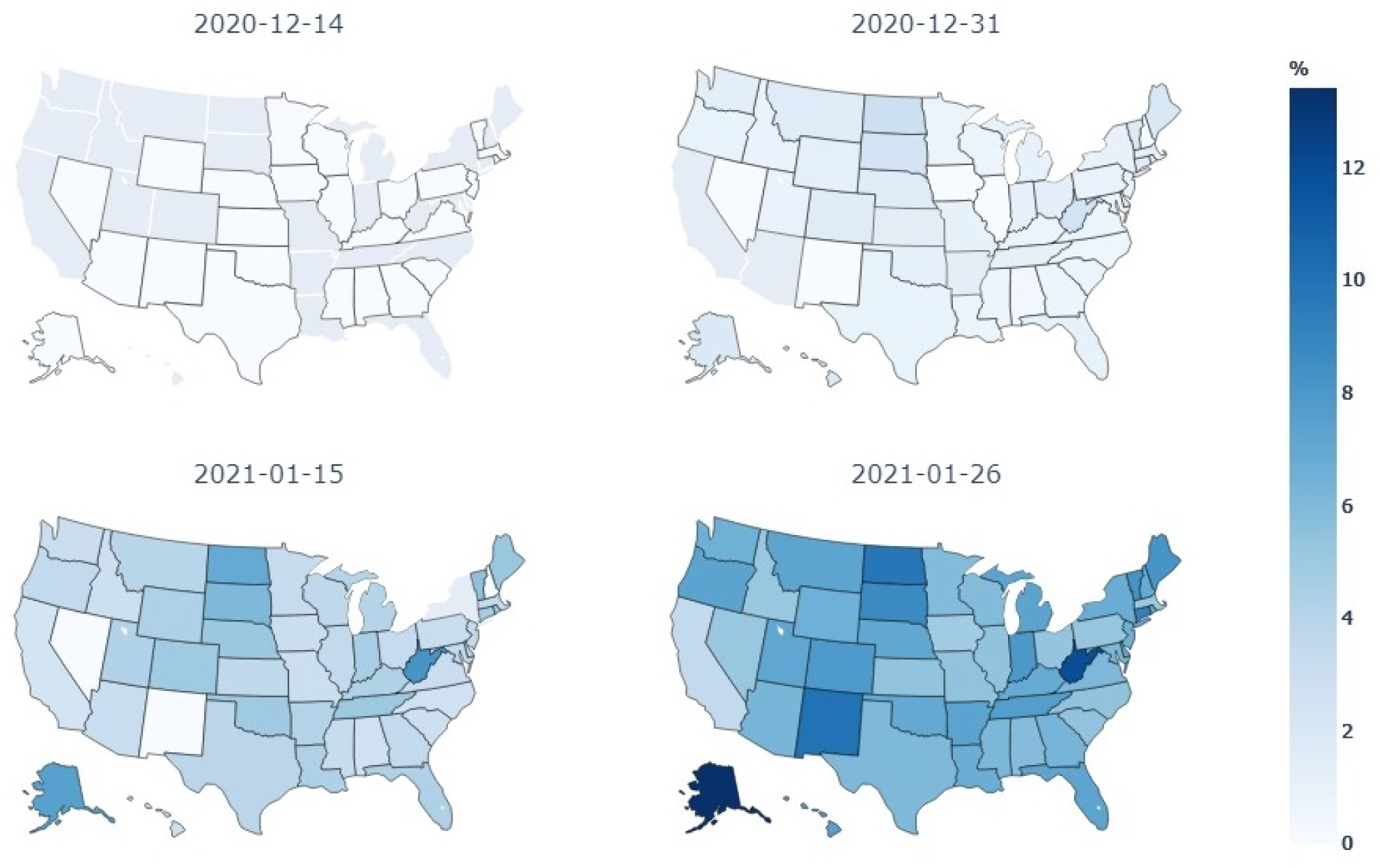
Percentage of the population who received at least 1 dose in the study period.

### Vaccination effectiveness

Figure 3 displays the effects of statewide COVID-19 vaccination on the state-level daily confirmed case growth rates based on the collected data. In addition, change in the daily growth rate percentage for the six post-periods and the four pre-event periods with respect to the reference period can also be visualized in Figure 3. The red vertical line in Figure 3 represents the reference period and the black square symbols describe the point estimates (along with their 95% confidence intervals) of the vaccination effect over different time periods. The figure shows that there were significant decreases in the daily COVID-19 case growth rate after the initial shots of the vaccine, with 0.124, 0.347, 0.345, 0.464, 0.490, and 0.756 percentage point declines in the 1-5, 6-10, 11-15, 16-20, 21-25, and 26 or more day post-periods after the start of vaccination.

**Figure 3:**
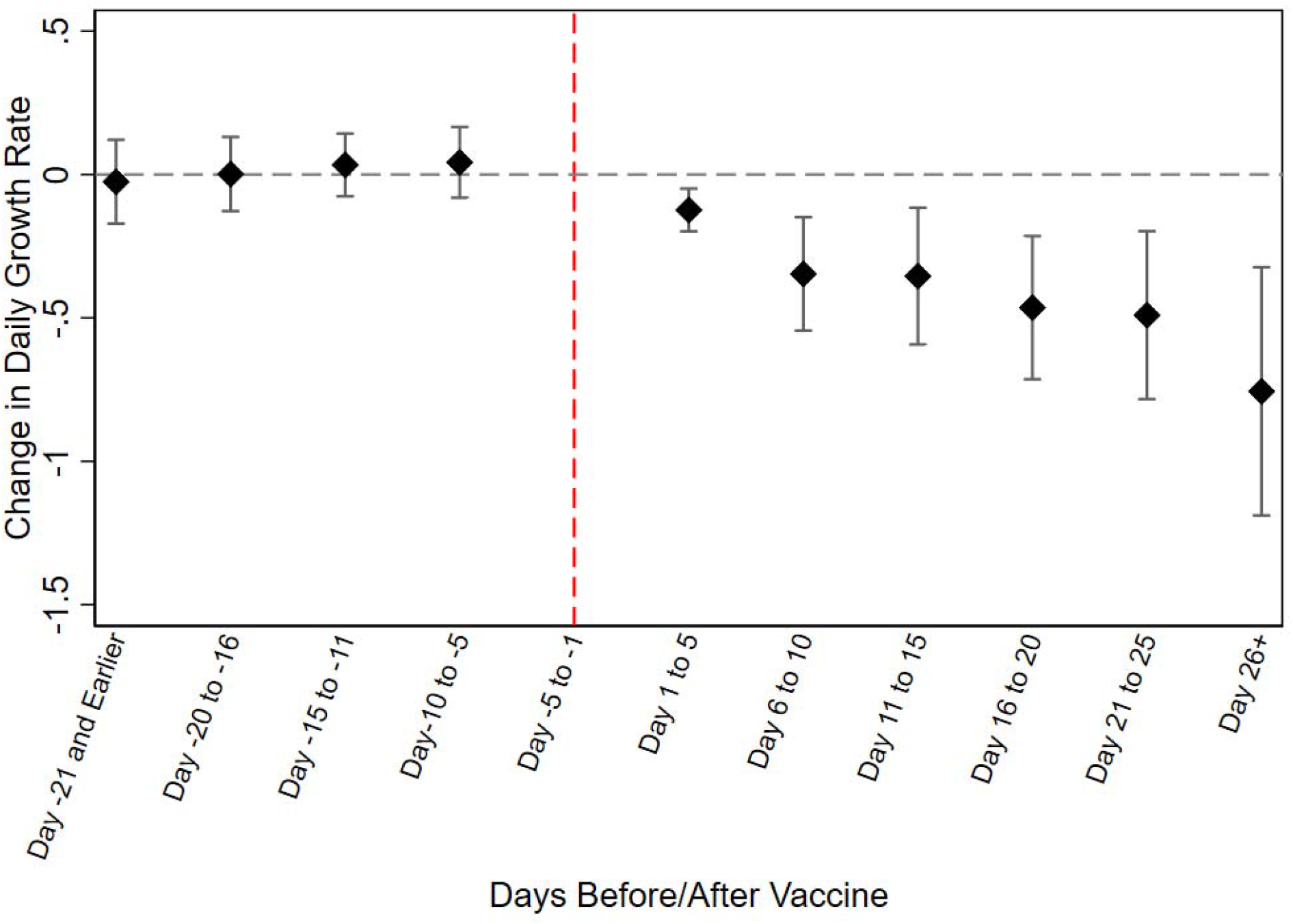
Event-study estimates of the effects of state vaccination on the daily state-level growth rate of COVID-19 confirmed cases.

### Robustness check

To verify the robustness of the estimates from the regression model, robustness checks were performed using different controls and samples. The main results column in Table 1 shows the effects of vaccination on the case growth rate with controlling for the Christmas holidays and pre-vaccination trends as explained previously. Again, this is the main regression we adopted, of which the average vaccine effects were plotted in Figure 3. Three other versions of the model were run—controlling for the Christmas holidays, controlling for the pre-vaccination trend, and without controlling for the Christmas holidays, as shown in Column 2, 3 and 4 respectively. These results showed similar effects on the case growth rate as seen with the main result. Columns 5-7 are robustness checks using the main regression specification but with some states excluded. Alabama was excluded in Column 5 because it has one of the lowest vaccination rates, while West Virginia was excluded in Column 6 because it has one of the highest vaccination rates. New Mexico, Wisconsin, Delaware, Tennessee, and Alaska were excluded from the test in Column 7 as these states represented a wide range in their percentage of distributed vaccines that have been administered (Adams et al., 2021). The results are robust in regard to these exclusions as they display similar results to what was seen in the main analysis.

**Table 1:** The administration date of the first COVID-19 vaccine shot in every state

| Date | States That Administered Their First Shot |
| --- | --- |
| 12/14/2020 | Arkansas, California, Colorado, Connecticut, Florida, Idaho, Indiana, Iowa, Kansas, Kentucky, Louisiana, Maryland, Michigan, Mississippi, Nebraska, Nevada, New York, North Carolina, North Dakota, Ohio, Pennsylvania, Rhode Island, South Dakota, Texas, West Virginia |
| 12/15/2020 | Alaska, Alabama, Arizona, Delaware, Georgia, Hawaii, Illinois, Maine, Massachusetts, Minnesota, Missouri, Montana, New Hampshire, New Jersey, New Mexico, Oklahoma, South Carolina, Utah, Virginia, Vermont, Washington, Wisconsin, Wyoming |
| 12/16/2020 | Oregon, Tennessee |

**Table 3:** The impact of vaccination on COVID-19 confirmed cases: Results from different models.

|  | (1)<br>Main results<br>(Controlling for<br>Christmas<br>holidays and<br>pre-vaccination<br>trend) | (2)<br>Controlling for<br>Christmas<br>holidays | (3)<br>Controlling<br>for pre-vaccination<br>trend | (4)<br>Without<br>controlling<br>holiday and<br>pre-vaccination<br>trend | (5)<br>Excluding<br>AL | (6)<br>Excluding<br>WV | (7)<br>Excluding<br>NM, WI, DE,<br>TN, AK |
| --- | --- | --- | --- | --- | --- | --- | --- |
| 21 or More<br>Days Before | -0.025 | 0.803*** | -0.025 | 0.802*** | -0.022 | -0.024 | -0.026 |
| 16 to 20 Days<br>Before | 0.001 | 0.303*** | 0.001 | 0.303*** | 0.003 | 0.001 | 0.015 |
| 11 to 15 Days<br>Before | 0.033 | 0.333*** | 0.034 | 0.333*** | 0.034 | 0.033 | 0.035 |
| 6 to 10 Days<br>Before | 0.042 | 0.269*** | 0.042 | 0.269*** | 0.043 | 0.043 | 0.036 |
| 1 to 5 Days<br>After | -0.124*** | -0.291*** | -0.124*** | -0.291*** | -0.137*** | -0.133*** | -0.107*** |
| 6 to 10 Days<br>After | -0.347*** | -0.409*** | -0.590*** | -0.650*** | -0.354*** | -0.392*** | -0.310*** |
| 11 to 15 Days<br>After | -0.354*** | -0.445*** | -0.649*** | -0.736*** | -0.360*** | -0.412*** | -0.275** |
| 16 to 20 Days<br>After | -0.464*** | -0.585*** | -0.482*** | -0.599*** | -0.473*** | -0.488*** | -0.431*** |
| 21 to 25 Days | -0.490*** | -0.625*** | -0.488*** | -0.621*** | -0.501*** | -0.514*** | -0.460*** |
| After |  |  |  |  |  |  |  |
| 26 or More Days After | -0.756*** | -0.951*** | -0.752*** | -0.943*** | -0.771*** | -0.755*** | -0.727*** |
| Stringency index | -0.002 | 0.014** | -0.002 | 0.014** | -0.002 | -0.002 | -0.001 |
| Test growth rate | 0.199 | -0.078 | 0.2 | -0.077 | 0.199 | 0.198 | 0.191 |
| Doses admin ratio | -0.069 | -0.05 | -0.07 | -0.052 | -0.066 | -0.067 | -0.077 |
| _cons | 0.464 | 0.532 | 0.464 | 0.53 | 0.436 | 0.464 | 0.438 |

## Discussion

Since the early stages of the pandemic, governments have distributed countless resources to support the development of the COVID-19 vaccine. Indeed, the vaccine has been claimed to be a vital tool in fighting the virus among other mitigation strategies and social distancing measures. However, the early administration of vaccines has been met with notable contention. Much public uncertainty can be attributed to a lack of quantitative evidence regarding how vaccination affects COVID-19 spread. This study is especially relevant as economies have started to reopen and social distancing compliance is declining, while cases are still at some of their highest totals. Vaccination efforts in the US commenced in mid to late December. Through the evaluation of certain periods past this initial adoption, our study showed that vaccination could significantly decrease the case growth rate in short-term, subsequent time periods.

Although vaccinations are generally effective in reducing the COVID-19 case growth rate, vaccination implementation and its effects on the case growth rate varied by state. The magnitude of decline in the case growth rate may be affected by factors that are heterogeneous among states, such as population density and other variables that have been associated with COVID-19. Additionally, inconsistency in implementation strategies can be seen in how well each state reached the public. For example, West Virginia increased vaccine administration dramatically throughout this analysis period. Successful vaccination in West Virginia, was partly attributed to the use of a network of pharmacies which facilitated local distribution (Anderson et al., 2021). Indeed, vaccination allocation may play a role in reaching different communities (Baumgartner et al., 2020). Additionally, as vaccination is a contentious issue and many people are skeptical of the vaccine due to a lack of quantitative information highlighting its effectiveness, many experts have recommended utilizing public information campaigns and media outlets to help spread information. These sources are vital in highlighting the safety and effectiveness of the vaccine to the public (Baumgartner et al., 2020; Wakefield et al., 2010). Making sure mitigation measures reach the public is an essential step in the ultimate goal of widespread vaccination and the fight against COVID-19. Other states can learn from these successful implementation strategies in order to reach a larger percentage of the public in later phases.

Our study contributes direct, empirical evidence, which highlights the effectiveness of the COVID-19 vaccination on the case growth rate. Overall, vaccination has been successful in decreasing the case growth rate during the early stages of vaccination despite the initial limited supply and slow rollout. Furthermore, the decreases increased in magnitude overtime. Although the vaccine is still in the early stages of administration. This information could be used to convince the public that vaccination is worthwhile and can help policymakers optimize implementation strategies.

Despite these promising results, this study was limited by the available data. For example, the real number of COVID-19 cases is likely much higher than the totals being reported (Beusekon, 2021). Future work may also investigate possible consequences of the vaccine using death case data which can better represent the severity of the virus. Similarly, the model used in this analysis failed to control for factors such as mask use and seasonality due to data availability. The vaccination approach and system will shift when more COVID-19 vaccines are approved by the FDA for regular distribution and supplies become more readily available (Means et al., 2020). Future research is warranted to estimate the reach, effectiveness, adoption, implementation, and maintenance of regular COVID-19 vaccination.

## Conclusion

This study found that the phased vaccination approach helped reduce the daily, state-level COVID-19 case growth rates in the US. Despite initially limited vaccine availability, the CDC guidelines regarding the phased vaccination approach has helped protect the most vulnerable from COVID-19. Even though the COVID-19 vaccine is still in the early stages of distribution, it has already been associated with significant decreases in the case growth rate. This is promising for the subsequent phases of vaccination. Overall, the results presented in this research have implications in regard to general public vaccine concerns. For example, our assertion that the COVID-19 vaccine is effective, even in the initial stage, may help persuade skeptical vaccine critics that this mitigation measure is worth it. This information is vital as states are reopening and social distancing measures are becoming less feasible both administratively and economically.

## Data Availability

https://github.com/stccenter/COVID-19-Data

